# Non-Communicable Disease Service Readiness in Nepal: A Further Analysis of Nepal Health Facility Survey- 2021

**DOI:** 10.1101/2023.02.07.23285512

**Authors:** Bikram Adhikari, Achyut Raj Pandey, Bipul Lamichhane, KC Saugat Pratap, Deepak Joshi, Shophika Regmi, Santosh Giri, Sushil Chandra Baral

## Abstract

**Objective:** To assess the readiness of public and private health facilities(HFs) in delivering Cardiovascular Diseases(CVDs), Diabetes Mellitus(DM), Chronic Respiratory Diseases(CRDs), and Mental Health(MH) services in Nepal.

**Methods:** We analyzed data on service readiness for CVDs, DM, CRDs, and MH from Nepal Health Facility Survey 2021 using Service Availability and Readiness Assessment manual of the World Health Organization. Readiness score was measured as the average availability of tracer items in percent, and facilities were considered “ready” for Non-Communicable Diseases (NCDs) management if scored ≥70 (out of 100). We performed weighted descriptive analysis, univariate and multivariable logistic regression to determine association of readiness of HFs with province, type of HFs, ecological region, quality assurance activities, external supervision, client’s opinion review, and frequency of HF meetings. The result of regression analysis are presented as odds ratio with 95% confidence interval(CI) and p-value.

**Results:** Of 1581 facilities offering any NCDs related services, 93.1%(95% CI: 90.9 to 94.7), 75.8%(95%CI: 72.4 to 78.8), 99.3%(95%CI: 98.3 to 99.7) and 26.0%(95%CI: 23.0 to 29.2) provide CVDs, DM, CRDs and MH-related services respectively. The overall readiness score for CVDs, DM, CRDs, and MH-related services were 38.1±15.4, 38.5±16.7, 32.6±14.7 and 24.0±23.1 respectively with readiness score lowest for the guidelines and staff training domain and highest for essential equipment and supplies domain. Peripheral public HFs were more likely to be ready to provide all NCDs-related services as compared to federal/provincial facilities. The HFs with external supervision in past 4 months were less likely to be ready to provide CRDs and DM related services and HFs reviewing client’s opinions were more likely to be ready to provide CRDs, CVDs and DM related services.

**Conclusion:** Readiness of HFs to provide CVDs, DM, CRDs, and MH-related services was sub-optimal in Nepal. It is recommended to reform policy to improve service readiness for NCDs.

**Strengths and limitations of this study:**

- Nationally representative sample of health facilities in Nepal, with coverage of all seven provinces and 77 districts
- Survey has adopted a highly standardized survey with the globally accepted research protocol. Variables for readiness analysis are based on standardized WHO’s SARA guideline and thus, findings are comparable to findings from other countries.
- Weighted analysis has been performed, which takes into account the complex sampling procedures and adjusts for non-response and disproportionate sampling.
- Since the survey was conducted while the COVID-19 pandemic was ongoing, there could be some impact of the pandemic on study results.

## Introduction

Non-communicable diseases (NCDs) have emerged as leading causes of premature mortality and Disability Adjusted Life Years (DALYs) in Nepal. Of the total 193,331 fatalities estimated in 2019, NCDs were responsible for 71.1% of deaths, while communicable, maternal, neonatal and nutritional conditions accounted for 21.1% of deaths, and the remaining 7.8% of deaths were due to injuries.^1^ In 2040, NCDs is projected to attribute 78.64% of total deaths in Nepal. ^1^ In 2019, cardiovascular diseases (CVD), Chronic respiratory disease and neoplasm were the top three leading causes of death in Nepal, attributing to approximately 24%, 21.1% and 11.2% of total deaths, respectively. Together, these three conditions are responsible for more than half of the total deaths in Nepal.^2^

Recognizing the gravity of the situation, Nepal adopted, contextualized and implemented the Package of Essential Noncommunicable Diseases (PEN) to screen, diagnose, treat, and refer major NCDs such as cardiovascular disease, diabetes, chronic obstructive pulmonary disease, cancer, and mental health at health posts, primary health care centers, and district hospitals.^3^ The PEN package has now been expanded to all 77 districts of Nepal.^4^ Moving a step further, the PEN Implementation Plan (2016-2020) was developed in accordance with the Multi-sectoral Action Plan for NCD Prevention and Control (2014-2020).^4^ Nepal Multi-Sectoral Action Plan for NCDs (2021-2025) focuses on creating high-impact, politically and socially acceptable, and potentially implementable interventions. The plan aims to reduce the burden of NCDs through the whole-of-government and whole-of-society approach. The action plan has an overarching target of reducing premature mortality from NCDs by 25% by 2025 and by one third by 2030.^4 5^ The NCD action plan is to achieve 80% availability of cheap basic technologies and necessary medications, including generics, needed to treat major NCDs in both public and private institutions. The multi-sectoral action plan involves medication therapy and counseling (including glycemic management) for 50% of eligible persons (defined as those aged 40 and older with a 10-year cardiovascular risk of more than 30%, including those with established cardiovascular disease).^5^

NCD services have been included in basic health care in Nepal although the service availability and preparedness remain very limited. ^6^ Apart from disease-specific interventions, Nepal Lancet NCDI poverty commission has pointed out the need for improving governance, strengthening health systems, and monitoring this extended group of priority NCDs. Commission also recommended that structured capacity-building programs for health service providers; promoting care packages, such as the PEN interventions for primary health care; increasing the availability of specialty services and personnel; and expanding progressive vertical programs providing free care for disease-specific areas could be useful in improving service availability and preparedness for NCDs.^7^

The increasing burden of NCDs in Nepal is often not matched with the availability of resources and appropriate healthcare response. There is a need to generate evidence to uncover gaps in NCD service readiness to facilitate evidence informed policy making to improve the service availability and uptake.^6 8^ We aim to determine services availability and readiness of health facilities in Nepal to provide services related to NCDs including CVDs, CRDs, DM, and MH using nationally representative data from Nepal Health Facility Survey (NHFS), 2021.

## Methods

### Study Design

We analyzed secondary data from the nationally representative cross-sectional survey, NHFS 2021 to assess the availability and readiness of health facilities to provide services related to NCDs namely, cardiovascular diseases (CVDs), chronic respiratory diseases (CRDs), diabetes mellitus (DM) and mental health (MH). Detailed information on survey objectives and methodology is published elsewhere.^9^

### Sample and Sampling

A stratified random sample of 1,633 health facilities out of 5,681 eligible health facilities were selected in 2021 NHFS. The process of sample size estimation and sampling procedures are explained in detail elsewhere.^9^ We analyzed data of 1487 facilities offering any NCDs services out of which 1387 facilities were offering CVDs services, 1474 were offering CRDs services, 1166 were offering DM services and 558 facilities were offering MH services.

### Data collection

Data collection for 2021 NHFS took place between January 27, 2021, and September 28, 2021, with a break for three months from May through July, due to the COVID-19 imposed lockdowns beginning on April 29, 2021. The 2021 NHFS included the use of four types of survey instruments: a) Facility Inventory Questionnaire, b) Health Provider Questionnaire, c) Exit Interview Questionnaires, and d) Observation protocols for antenatal care, family planning services, care for sick children, and labor and delivery. We used data from the first instrument, Facility Inventory Questionnaire.

### Patient and public involvement

This article is prepared analyzing secondary data sources. There were no patient and public involvement in the design, conduct and reporting of our research.

### Outcome variables and measurement

The variables for services availability and readiness of health facilities to provide NCDs-related facilities were selected based on the WHO Service Availability and Readiness (SARA) manual. [8]

#### Services Readiness

The service readiness of health facilities was measured based on the availability and functioning of items categorized under three domains-staff and guidelines, essential equipment, and supplies, medicine and commodities and diagnostics. The list of tracer items of each domain for CVDs, CRDs, DM and MH are presented in the **supplementary file 1**. The readiness score of health facilities to provide services on CVDs, CRDs DM and MH were calculated using the Service Availability and Readiness Assessment manual of the World Health Organization.[8] The items in each domain were re-coded as binary variables, taking “1” for the presence of the item and “0” for the absence of the item in the facility. The mean score for each domain was computed by adding items followed by dividing by the number of items and multiplying by 100. The average of the score from the three domains was the readiness score. A cutoff of 70 was considered on the overall score to classify the readiness of the facilities towards NCDs-related services. A facility with an overall score of more than or equal to 70 was considered as ready to manage NCDs. ^10–12^

### Independent variables

The independent variables included setting (rural/urban), ecological region (Hill/Mountain / Terai), province (Province-1/ Madhesh / Bagmati / Gandaki /Lumbini / Karnali /Sudurpaschim), type of facility (federal or provincial hospital/peripheral facilities/private hospital), presence of external supervision in past four months (present/absent), quality assurance activities (performed /not performed) and frequency of health facility meeting (none/sometimes/monthly).

The classification of setting into rural and urban was based on the type of municipalities in which the health facilities are located.^9^ The type of facilities were classified into federal or provincial hospital, peripheral facilities and private hospitals where peripheral facilities comprised of hospitals, health posts and primary health care centers. The facility that received any external supervision/monitoring from the federal, provincial or municipal level in the past four months was considered to have external supervision.^9^ Facility which routinely carries out quality assurance activities and had documentation of a recent quality assurance activity including report or minutes of a quality assurance meeting, a supervisory checklist, a mortality review, or an audit of records or registers were considered to have performed quality assurance activities. ^9^ The frequency of health facilities stating “no” for routine management/administrative meetings were classified as “None”. Similarly, those stating “monthly or more often” were classified as “Monthly” and those stating “irregular or every 2-6 months” were classified as “sometimes”.^9^ The facility with the system for determining client opinion, procedure for reviewing client opinion, and report of a recent review of client opinion were considered to have performed review of client’s opinion.^9^

### Statistical Analysis

We used R version 4.2.0 and RStudio ^13 14^ for statistical analysis. We used “survey” package ^15^ and performed a weighted analysis to account for the complex survey design of 2021 NHFS. We summarized continuous variables with weighted and unweighted mean, standard deviation (SD), median and interquartile range (IQR) whereas categorical variables were summarized with both weighted and weighted frequency, percent, and 95 percent confidence interval (95% CI) around percent. We employed univariate and multivariate weighted logistic regression analysis to determine the association of the readiness of health facilities to CVDs, CRDs, DM and MH-related services with independent variables including setting, ecological region, province, type of facility, external supervision, quality assurance activities and health facility meeting. A p-value of less than 0.05 is considered statistically significant.

### Ethical approval

The original 2021 NHFS survey was approved by the Institutional Review Board of ICF International, USA, and by the Nepal Health Research Council (NHRC), Nepal.^9^

## Results

Of the total facilities offering any NCDs (CVDs, CRDs, DM or MH) related services, 46.6% (95% CI: 42.9 to 50.4) were from rural areas. Half of the health facilities with any NCDs related services were from hill region (52.7%; 95% CI: 48.9 to 56.5) followed by terai region (34.4%; 95% CI: 30.8 to 38.2). The health facilities from Bagmati province was the highest of all accounting for 20.5% (95% CI: 17.6 to 23.7) followed by Madhesh province (16.1%; 95% CI:13.1 to 19.7), province-1 (15.7%; 95% CI:13.0 to 18.8) and Lumbini (15.7%; 95% CI:13.2 to 18.6) The quality assurance activities for at least once a year was performed in 23.9% (95% CI: 20.7 to 27.4), external supervision in past 4 months was present in 66.4 (95% CI: 62.8, 69.8). Review of client’s opinion in 3.8% (95% CI: 2.7 to 5.3) and monthly health facility meeting was carried out in 65.0% (95% CI: 61.4 to 68.4) of the facilities offering any NCDs related services. (*Table-1*)

**Table 1:** Characteristics of facilities offering any NCDs related services

| Characteristics | Unweighted (N=1487) | Weighted (N=1518) |  |
| --- | --- | --- | --- |
|  | n(%) | n(%) | 95%CI |
| **Location** |  |  |  |
| Urban | 953 (64.1) | 810(53.4) | 49.6 to 57.1 |
| Rural | 534 (35.9) | 708 (46.6) | 42.9 to 50.4 |
| **Ecological Region** |  |  |  |
| Hill | 797 (53.6) | 801 (52.7) | 48.9 to 56.5 |
| Mountain | 178 (12.0) | 196 (12.9) | 10.6 to 15.5 |
| Terai | 512 (34.4) | 522 (34.4) | 30.8 to 38.2 |
| **Province** |  |  |  |
| Province-1 | 227 (15.3) | 238 (15.7) | 13.0 to 18.8 |
| Madhesh | 170 (11.4) | 245 (16.1) | 13.1 to 19.7 |
| Bagmati | 295 (19.8) | 311(20.5) | 17.6 to 23.7 |
| Gandaki | 220 (14.8) | 197 (13.0) | 10.8 to 15.5 |
| Lumbini | 238 (16.0) | 238 (15.7) | 13.2 to 18.6 |
| Karnali | 143 (9.6) | 119 (7.8) | 6.4 to 9.6 |
| Sudurpaschim | 194 (13.0) | 169 (11.1) | 9.3 to 13.2 |
| **Ownership** |  |  |  |
| Private hospitals | 255 (17.1) | 113 (7.5) | 6.2 to 8.9 |
| Public health facilities | 1,232 (82.9) | 1,405 (92.5) | 91.1 to 93.8 |
| **Type of facility** |  |  |  |
| Federal/Provincial hospital | 97 (6.5) | 27 (1.8) | 1.5 to 2.2 |
| Local hospital/PHCC/peripheral | 1,135 (76.3) | 1,378 (90.7) | 89.2 to 92.0 |
| Private Hospital | 255 (17.1) | 113 (7.5) | 6.2 to 8.9 |
| **Quality assurance activities** |  |  |  |
| Not Performed | 1,189 (80.0) | 1,155 (76.1) | 72.6 to 79.3 |
| Performed | 298 (20.0) | 363 (23.9) | 20.7 to 27.4 |
| **External Supervision** |  |  |  |
| No | 564 (37.9) | 510 (33.6) | 30.2 to 37.2 |
| Yes | 923 (62.1) | 1008 (66.4) | 62.8 to 69.8 |
| **Review client's opinion** |  |  |  |
| Not reviewed | 1,400 (94.1) | 1461 (96.2) | 94.7 to 97.3 |
| Reviewed | 87 (5.9) | 57 (3.8) | 2.7 to 5.3 |
| **Frequency of health facility meeting** |  |  |  |
| None | 264 (17.8) | 225 (14.8) | 12.5 to 17.5 |
| Sometimes | 303 (20.4) | 306 (20.2) | 17.4 to 23.3 |
| Monthly | 920 (61.9) | 987 (65.0) | 61.4 to 68.4 |
| **Total Health workforce<sup>1</sup>** | 40.9(122.8);<br>7.0(3.0,20.0) | 18.7±73.5; 6.0 (5.0, 9.0) |  |
| **Services availability** |  |  |  |
| CVDs | 1387 (93.3) | 1411 (93.1) | 90.9 to 94.7 |
| DM | 1166 (78.4) | 1149 (75.8) | 72.4 to 78.8 |
| CRDs | 1474 (99.1) | 1507 (99.3) | 98.3 to 99.7 |
| MH | 558 (37.5) | 394 (26.0) | 23.0 to 29.2 |
| <sup>1</sup> Mean±SD; Median (IQR)<br>CVD: Cardiovascular Disease; DM: Diabetes Mellitus; CRDs: Chronic Respiratory Disease; MH: Mental Health |  |  |  |

Table 2 presents the overall readiness score of health facilities for CVDs, CRDs, DM, and MH-related services (Mean±SD) were 38.1±15.4, 32.6±14.7, 38.5±16.7, and 24.0±23.1 respectively. The readiness score for guidelines and staff training domain for NCDs related services ranged from 12.1 to 17.6 and was lowest in all sets of facilities offering different NCDs related services whereas the readiness score for essential equipment and supplies domain ranged from 49.1 to 76.4 and was highest in three sets of facilities offering CVDs, DM and CRDs related services. The percent of total facilities which are ready to provide CVDs, CRDs, DM and MH-related services were 3.8% (95% CI: 5.3 to 2.8), 2.3% (95% CI: 1.6 to 3.3), 3.7% (95% CI: 2.6 to 5.1) and 3.3% (95% CI: 1.8 to 6.1) respectively.

**Table 2:**
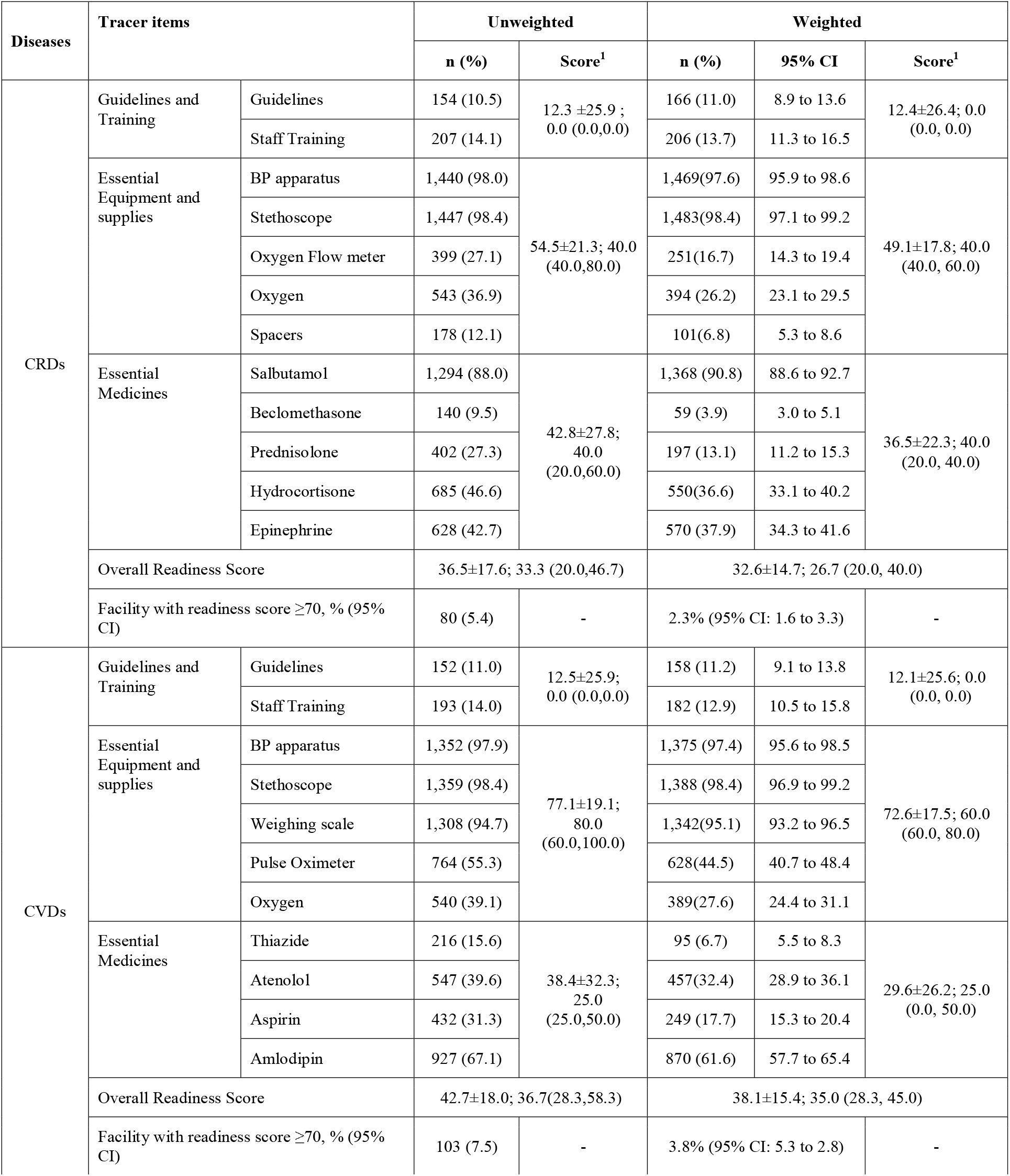
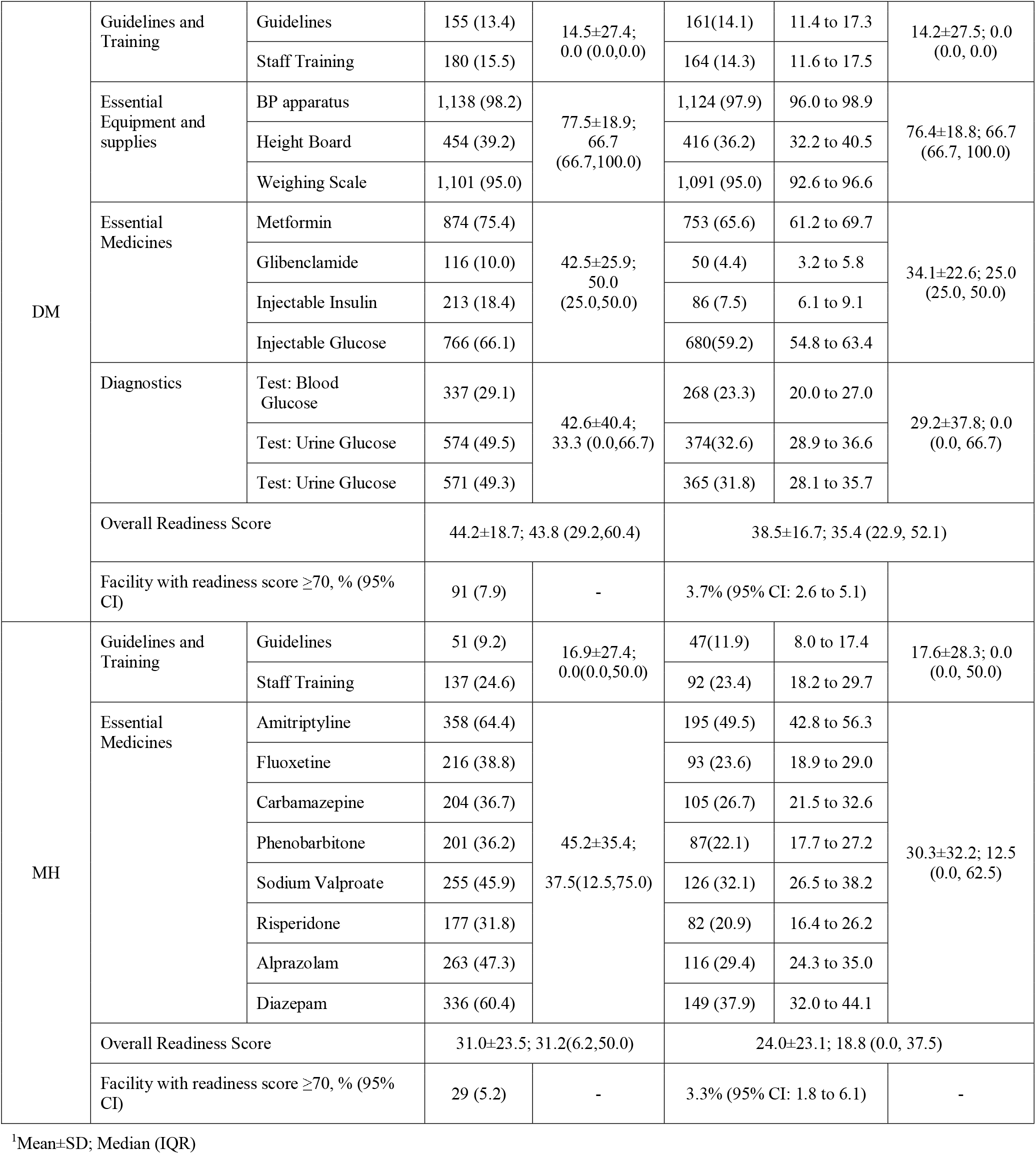
Readiness of facilities for services related to NCDs.

Figure 1 shows the readiness of facilities to provide NCDs services grouped by district. Facilities are represented by red points for readiness scores ranging from 0 to 20 and white points for readiness scores ranging from 80 to 100, while districts with readiness scores from 0 to 20 are represented by yellow color and with readiness scores from 80 to 100 in blue. These legends are consistent across all the sub-figures A through D.

**Figure 1:**
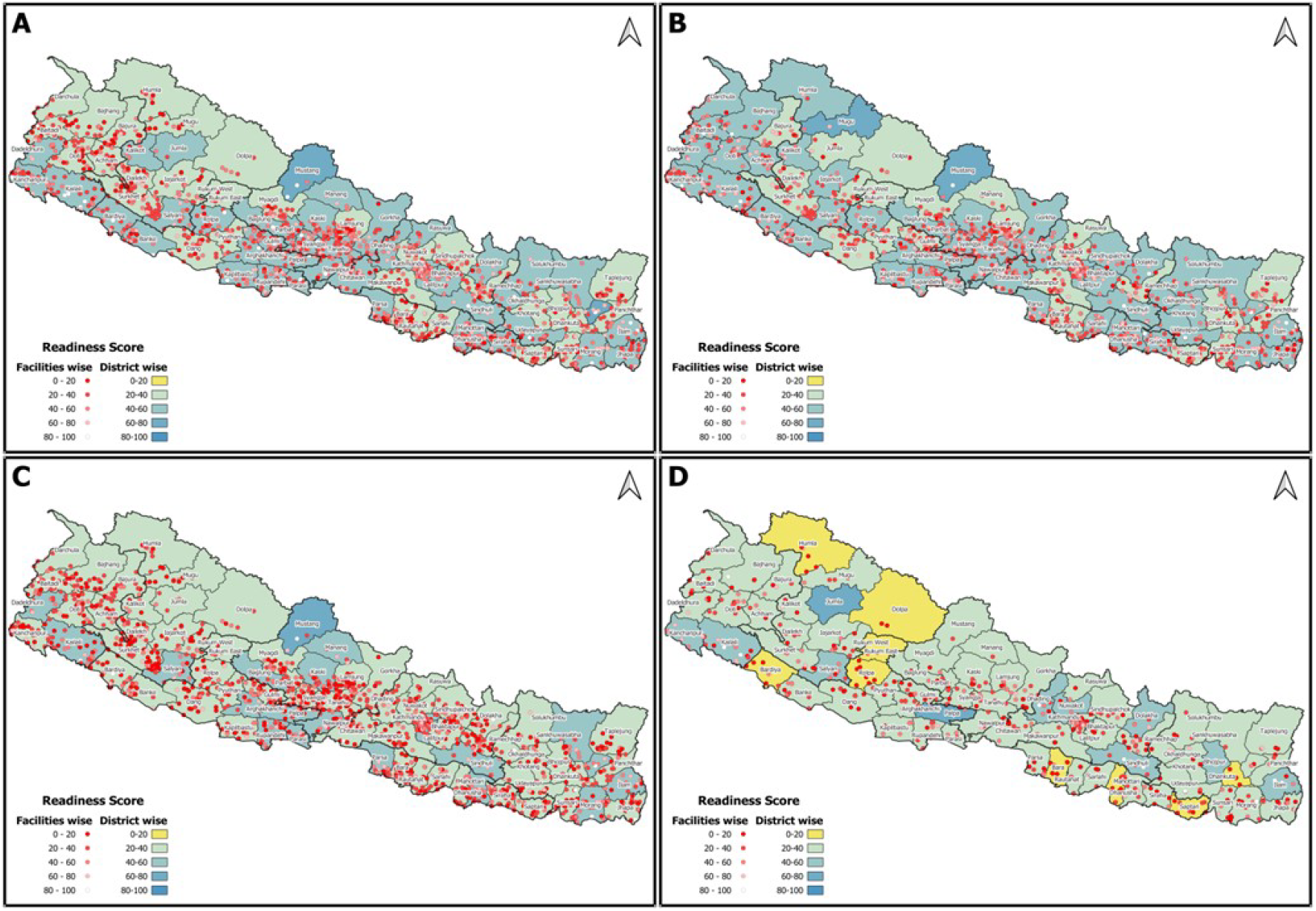
Facilities and district-wise readiness score of facilities to provide services related to NCDs (A: Readiness of facilities to provide CVD-related services; B: Readiness of facilities to provide DM related services; C. Readiness of facilities to provide CRDs related services; D: Readiness of facilities to provide MH related services)

Gardner-Altman estimation plot to compare the readiness score of health facilities for providing different NCD-related services by province showed the readiness score of facilities vary by province. (**Supplementary file 2**)

Table 3 and 4 presents the factors associated with services readiness of health facilities to provide NCDs-related services. In univariate analysis, service readiness of health facilities to provide chronic respiratory disease were significantly associated with the type of facility and presence of supervision and revision of client opinion. Similarly, the readiness of health facilities for CVDs were significantly associated with the revision of client opinion and type of health facility. The readiness of facilities for diabetes mellitus was significantly associated with the type of facility, presence of quality assurance performed at least once a year, presence of external supervision, and revision of client opinion. The readiness of facilities for mental health services were associated with the type of facility.

**Table 3:** Factors associated with readiness of health facilities to provide NCDs related services (unadjusted)

| Characteristics | Chronic Respiratory Disease related services |  | Cardiovascular Disease related services |  | Diabetes Mellitus related services |  | Mental Health related services |  |
| --- | --- | --- | --- | --- | --- | --- | --- | --- |
|  | COR (95% CI) | p | COR (95% CI) | p | COR (95% CI) | p | COR (95% CI) | p |
| **Location** |  |  |  |  |  |  |  |  |
| Urban | Ref |  | Ref |  | Ref |  | Ref |  |
| Rural | 0.70 (0.31 to 1.57) | 0.39 | 1.03 (0.52 to 2.03) | 0.93 | 0.49 (0.21 to 1.11) | 0.09 | 0.83 (0.16 to 4.17) | 0.80 |
| **Ecological region** |  |  |  |  |  |  |  |  |
| Hill | Ref |  | Ref |  | Ref |  | Ref |  |
| Mountain | 0.99 (0.29 to 3.33) | 0.98 | **0.22 (0.09 to 0.57)** | **0.00** | 0.74 (0.32 to 1.74) | 0.50 | 1.18 (0.13 to 10.78) | 0.90 |
| Terai | 0.78 (0.37 to 1.64) | 0.51 | 0.80 (0.40 to 1.60) | 0.52 | 1.42 (0.68 to 2.96) | 0.40 | 0.79 (0.26 to 2.41) | 0.70 |
| **Province** |  |  |  |  |  |  |  |  |
| Province-1 | Ref |  | Ref |  | Ref |  | Ref |  |
| Madhesh | 0.50 (0.19 to 1.33) | 0.17 | **0.25 (0.09 to 0.73)** | **0.01** | 0.56 (0.23 to 1.37) | 0.20 | 0.61 (0.11 to 3.53) | 0.60 |
| Bagmati | 1.65 (0.64 to 4.26) | 0.30 | 0.97 (0.33 to 2.89) | 0.96 | 2.09 (0.81 to 5.39) | 0.13 | 2.24 (0.46 to 10.76) | 0.30 |
| Gandaki | 2.46 (0.84 to 7.16) | 0.10 | 1.15 (0.38 to 3.43) | 0.80 | 1.50 (0.51 to 4.37) | 0.50 | 0.48 (0.09 to 2.72) | 0.40 |
| Lumbini | 1.51 (0.47 to 4.79) | 0.49 | 1.04 (0.34 to 3.15) | 0.95 | 1.36 (0.48 to 3.87) | 0.60 | 0.43 (0.09 to 2.00) | 0.30 |
| Karnali | 0.26 (0.06 to 1.19) | 0.08 | 0.49 (0.10 to 2.42) | 0.38 | 0.46 (0.13 to 1.64) | 0.20 | 0.59 (0.10 to 3.38) | 0.60 |
| Sudurpaschim | 1.50 (0.68 to 3.31) | 0.31 | 0.79 (0.27 to 2.36) | 0.68 | **2.49 (0.97 to 6.38)** | **0.06** | 2.98 (0.61 to 14.59) | 0.20 |
| **Type of facility** |  |  |  |  |  |  |  |  |
| Federal/Provincial hospitals | Ref |  | Ref |  | Ref |  | Ref |  |
| Peripheral health facilities | **0.06 (0.03 to 0.13)** | **<0.001** | **0.15 (0.08 to 0.31)** | **<0.001** | **0.08 (0.04 to 0.17)** | **<0.001** | **0.17 (0.05 to 0.58)** | **0.01** |
| Private hospital | 0.36 (0.14 to 0.89) | 0.03 | 0.64 (0.29 to 1.42) | 0.30 | **0.39 (0.18 to 0.85)** | **0.02** | 0.21 (0.08 to 0.54) | 0.00 |
| **Quality assurance activities** |  |  |  |  |  |  |  |  |
| Not performed | Ref |  | Ref |  | Ref |  | Ref |  |
| Performed | 1.03 (0.56 to 1.88) | 0.93 | 0.81 (0.44 to 1.49) | 0.50 | **2.18 (1.08 to 4.42)** | **0.03** | 0.56 (0.20 to 1.57) | 0.30 |
| **External supervision** |  |  |  |  |  |  |  |  |
| No | Ref |  | Ref |  | Ref |  | Ref |  |
| Yes | **2.68 (1.43 to 5.04)** | **0.00** | 1.57 (0.76 to 3.22) | 0.22 | **2.25 (1.23 to 4.10)** | **0.01** | 0.83 (0.23 to 2.96) | 0.80 |
| **Frequency of health facility meeting** |  |  |  |  |  |  |  |  |
| None | Ref |  | Ref |  | Ref |  | Ref |  |
| Sometimes | **0.29 (0.10 to 0.85)** | **0.02** | 0.84 (0.17 to 4.10) | 0.83 | **3.53 (1.13 to 11.03)** | **0.03** | 0.94 (0.17 to 5.21) | 0.94 |
| Monthly | 0.45 (0.16 to 1.25) | 0.13 | 2.01 (0.54 to 7.44) | 0.30 | **3.55 (1.40 to 9.04)** | **0.01** | 2.25 (0.43 to 11.87) | 0.34 |
| **Review of client's opinion** |  |  |  |  |  |  |  |  |
| Not reviewed | Ref |  | Ref |  | Ref |  | Ref |  |
| Reviewed | **5.05 (1.79 to 14.28)** | **0.002** | **4.87 (2.14 to 11.06)** | **<0.001** | **4.84 (2.09 to 11.21)** | **<0.001** | 2.88 (0.92 to 9.02) | 0.07 |

*COR: Crude Odds ratio; CI: Confidence Interval; Ref: Reference group*
*Peripheral health facilities include local hospital, health posts and primary health care centers*

**Table 4:** Factors associated with readiness of health facilities to provide NCDs-related services (adjusted)

| Characteristics | Chronic Respiratory Disease related services |  | Cardiovascular Disease related services |  | Diabetes Mellitus related services |  | Mental Health related services |  |
| --- | --- | --- | --- | --- | --- | --- | --- | --- |
|  | AOR (95% CI) | p | AOR (95% CI) | p | AOR (95% CI) | p | AOR (95% CI) | p |
| **Location** |  |  |  |  |  |  |  |  |
| Urban | Ref |  | Ref |  | Ref |  | Ref |  |
| Rural | 1.55 (0.74 to 3.21) | 0.24 | **2.04 (1.02 to 4.09)** | **0.045** | 1.28 (0.50 to 3.23) | 0.61 | 1.66 (0.12 to 23.80) | 0.71 |
| **Ecological region** |  |  |  |  |  |  |  |  |
| Hill | Ref |  | Ref |  | Ref |  | Ref |  |
| Mountain | 1.45 (0.39 to 5.47) | 0.58 | **0.24 (0.09 to 0.65)** | **0.005** | 0.82 (0.34 to 1.96) | 0.65 | 1.11 (0.08 to 15.24) | 0.94 |
| Terai | 1.15 (0.39 to 3.37) | 0.80 | 1.18 (0.38 to 3.70) | 0.78 | 2.74 (0.91 to 8.22) | 0.072 | 1.45 (0.47 to 4.46) | 0.52 |
| **Province** |  |  |  |  |  |  |  |  |
| Province-1 | Ref |  | Ref |  | Ref |  | Ref |  |
| Madhesh | 0.63 (0.19 to 2.09) | 0.45 | **0.24 (0.07 to 0.78)** | **0.018** | 0.43 (0.16 to 1.18) | 0.10 | 0.71 (0.10 to 5.20) | 0.74 |
| Bagmati | 1.58 (0.59 to 4.21) | 0.36 | 0.9 (0.23 to 3.45) | 0.87 | 2.39 (0.71 to 8.08) | 0.16 | 2.94 (0.41 to 20.95) | 0.28 |
| Gandaki | 2.81 (0.83 to 9.53) | 0.10 | 1.28 (0.36 to 4.54) | 0.70 | 2.75 (0.82 to 9.17) | 0.10 | 0.57 (0.10 to 3.23) | 0.52 |
| Lumbini | 1.66 (0.44 to 6.17) | 0.45 | 0.89 (0.27 to 2.99) | 0.86 | 1.22 (0.43 to 3.44) | 0.71 | 0.43 (0.07 to 2.70) | 0.36 |
| Karnali | 0.32 (0.07 to 1.51) | 0.15 | 0.76 (0.13 to 4.46) | 0.76 | 0.89 (0.21 to 3.82) | 0.87 | 0.78 (0.11 to 5.38) | 0.80 |
| Sudurpaschim | 2.25 (0.97 to 5.20) | 0.06 | 1.05 (0.32 to 3.42) | 0.94 | **3.31 (1.23 to 8.87)** | **0.02** | 3.71 (0.80 to 17.08) | 0.093 |
| **Type of facility** |  |  |  |  |  |  |  |  |
| Federal/Provincial hospital | Ref |  | Ref |  | Ref |  | Ref |  |
| Peripheral health facilities | **0.04 (0.02 to 0.09)** | **<0.001** | **0.12 (0.05 to 0.28)** | **<0.001** | **0.08 (0.04 to 0.18)** | **<0.001** | **0.17 (0.03 to 0.95)** | **0.04** |
| Private hospitals | **0.37 (0.16 to 0.87)** | **0.02** | 0.56 (0.24 to 1.31) | 0.18 | 0.42 (0.16 to 1.11) | 0.079 | **0.14 (0.04 to 0.55)** | **0.01** |
| **Quality assurance activities** |  |  |  |  |  |  |  |  |
| Not Performed | Ref |  | Ref |  | Ref |  | Ref |  |
| Performed | 0.86 (0.45 to 1.64) | 0.66 | 0.64 (0.33 to 1.25) | 0.19 | 2.06 (0.98 to 4.35) | 0.06 | 0.45 (0.16 to 1.29) | 0.14 |
| **External supervision** |  |  |  |  |  |  |  |  |
| No | Ref |  | Ref |  | Ref |  | Ref |  |
| Yes | **3.43 (1.64 to 7.20)** | **0.00** | 1.59 (0.69 to 3.66) | 0.27 | **2.56 (1.29 to 5.08)** | **0.01** | 0.85 (0.19 to 3.87) | 0.83 |
| **Frequency of health facility meeting** |  |  |  |  |  |  |  |  |
| None | Ref |  | Ref |  | Ref |  | Ref |  |
| Sometimes | **0.27 (0.10 to 0.76)** | **0.01** | 0.78 (0.15 to 3.99) | 0.76 | 2.77 (0.76 to 10.01) | 0.12 | 0.76 (0.13 to 4.54) | 0.76 |
| Monthly | **0.25 (0.09 to 0.65)** | **0.00** | 1.51 (0.38 to 5.90) | 0.56 | 1.85 (0.69 to 4.92) | 0.22 | 1.84 (0.29 to 11.83) | 0.52 |
| **Review of client's opinion** |  |  |  |  |  |  |  |  |
| Not Reviewed | Ref |  | Ref |  | Ref |  | Ref |  |
| Reviewed | 2.60 (0.91 to 7.44) | 0.07 | **2.68 (1.26 to 5.70)** | **0.01** | 1.99 (0.77 to 5.14) | 0.16 | 3.15 (0.97 to 10.19) | 0.06 |

*AOR: Adjusted Odds ratio; CI: Confidence Interval; Ref: reference group*
*Peripheral health facilities include local hospital, health posts and primary health care centers.*

In the adjusted multivariable analysis, the odds of being ready for chronic respiratory disease-related services were 0.04 (95% CI: 0.02 to 0.09) times in the peripheral public health facilities and 0.37 (95% CI: 0.16 to 0.87) times in facilities compared to federal/provincial facilities, and 3.43 (1.64 to 7.20) times in facilities with external supervision in past 4 months compared to facilities without external supervision in past 4 months after adjusting for other variables. The odds of being ready for CVDs related services were 2.04 (95% CI: 1.02 to 4.09) times in rural areas compared to urban, 0.24 (95% CI: 0.09 to 0.65) times in mountain compared to the hill, 0.24 (95% CI: 0.07 to 0.78) in Madhesh compared to Province-1, 0.12 (95% CI: 0.05 to 0.28) times in peripheral public health facilities compared to federal/provincial hospital and 2.68 (95% CI: 1.26 to 5.70) times in facilities reviewing client’s opinions compared those facilities that did not review client’s opinions. The odds of readiness of health facilities towards diabetes were 3.31 (95% CI: 1.23 to 8.87) times higher in Sudurpaschim compared to Province-1, 92% [AOR=0.08 (95% CI: 0.04 to 0.18)] less in peripherals compared to federal/provincial hospitals, and 2.56 (95% CI: 1.29 to 5.08) times in facilities with external supervision in past 4 months. Similarly, the odds of being ready for mental health services were 83% less in private hospitals [AOR=0.17 (95% CI: 0.03 to 0.95)] and 84% less in peripheral public health facilities [AOR=0.14 (95% CI: 0.04 to 0.55)] compared to federal/provincial hospitals.

## Discussion

This study aimed to determine the readiness of the health facilities to provide services related to NCDs including cardiovascular diseases, chronic respiratory disease, diabetes mellitus, and mental health in Nepal from a nationally representative sample of health facilities from the Nepal Health Facility Survey 2021. The overall median readiness score of the health facilities to provide CRDs, CVDs, diabetes, and mental health-related services were 26.7, 35.0, 35.4 and 18.8 respectively with the readiness score for guidelines and training domain being the lowest and the readiness score for essential equipment and supplies being the highest in each disease. Very few of the facilities offering respective NCDs-related services have readiness scores greater than 70 accounting for 3.8% for CVDs, 3.7% for diabetes, 3.3% for mental health, and 2.3% for chronic respiratory disease-related services. Peripheral public health facilities were more likely to be ready to offer all NCD-related services, compared to federal and provincial facilities. Facilities that received external supervision in the past 4 months were less likely to be ready to provide CRD and DM services, while facilities that considered client feedback were more likely to be ready to offer CRD, CVD, and DM services. The results of the study identified areas for improvement in the management of NCDs as well as strengths and shortcomings in the current healthcare system of Nepal.

A similar analysis from 2015 health facility survey showed the median readiness score of facilities to provide CVDs, CRDs, and DM to be 18.8, 11.3, and 26.4 respectively.^16^ This finding from 2015 was comparatively lower compared to the present study. One of the factors for the increase in the readiness score of health facilities from 2015 to 2021 could be due to the roll out and expansion of the Package of Essential NCDs (PEN). PEN aims to detect, diagnose, treat, and refer individuals with CVDs, Chronic Obstructive Pulmonary diseases, cancer, DM, and MH issues at health posts, primary health care centers, and district hospitals in order to promote early detection and management of chronic diseases within the community. ^3^ In October 2016, PEN was introduced in two pilot districts, Illam and Kailali which was later expanded to all 77 districts in Nepal^4^. In addition, the National mental health strategy and action plan was launched in 2021 which can further improve the preparedness of health system to deliver mental health services in future. ^4 17^

In our study, the availability of guidelines and staff training was one of the most problematic areas that needs intervention. A prior study on CVDs highlighted a lack of national guidelines and protocols for treating CVDs as a significant obstacle to providing evidence-based treatment. ^18^ The other study on DM suggested that there is a significant shortfall in the implementation of existing policies, plans, strategies, and programs aimed at addressing diabetes, with a lack of clarity on how they should be implemented. ^19^ This evidence suggests not only the need for formulation of evidence-informed guidelines and policies but also the appropriate communication and supply so facilities under all tiers of government have a clear understanding of the policy documents. These areas should be improved and addressed concurrently as they have been demonstrated to be cost-efficient in terms of healthcare delivery.^20^

In tune with our findings, previous studies have also shown disparities in the availability of healthcare resources for the prevention and control of NCDs between different levels of healthcare, types of facilities, and regional settings.^21^ Our study found that there was a significant lack of essential medicines and commodities for NCDs in public and rural facilities which was also reported in other studies ^22 23^. The shortages of essential medicines and commodities were often accompanied by shortages/lack of training of staff, which further hindered access to proper medical care for patients; which has also been the case for a study done in Nepal using the 2015 health facility survey data.^16^ This is a cause for concern as it can negatively impact the health outcomes of individuals suffering from NCDs.^24^ It is crucial to stress the relationship between the availability of drugs and supplies, and the training of health care professionals. For instance, even if trained personnel were available to provide services, a lack of drugs and supplies will prevent the health professional from providing quality healthcare, and the other way around.^25^ Therefore, there is an urgent need to address the scarcity of both trained personnel and medicines, since doing so might assist to enhance management, which in turn would lead to an increase in the supply and availability of medications. Our study revealed that facilities with external supervision had significantly higher preparedness scores for diabetes mellitus and chronic respiratory diseases. According to a study, external supervision mechanisms in healthcare facilities are essential in facilitating the overall management process and improving the effectiveness of the facility. Such supervision enables information sharing and performance review which is pivotal in streamlining the facility’s management process and enhancing its efficiency. ^26^

Within South Asian regions, differences regarding the lack of trained personnel, availability of essential medicines and commodities and guidelines in service-specific readiness have also been documented. ^27^ The region’s progress in the management and prevention of NCDs has been hampered by the widespread absence of key resources. According to a recent report by the WHO, most countries, particularly low- and middle-income countries (LMICs), failed to achieve the global targets set for noncommunicable disease (NCD) progress in 2020. This report, which evaluated data from 194 countries, highlights the pressing need for increased global efforts in NCD prevention and control.^28^

Alongside the issues discussed, Nepal’s health system has the potential to effectively address NCDs. Nepal has implemented policies and strategies, developed treatment guidelines and protocols, an essential drug list, a multisectoral plan for NCD prevention, surveillance and prevention strategic planning, and an action plan for NCDs. These findings suggest that Nepal should strengthen and orient health systems for the prevention and control of NCDs and strengthen supervision and monitoring as aligned with the action plan for the prevention and control of NCDs. ^17^ The disparities identified across various diseases and healthcare types and levels, as well as the noticeable differences in availability between urban and rural areas, along with a lack of basic medicines and supplies, underline the importance of an all-inclusive approach to upgrading healthcare facilities’ ability to deliver successful NCD interventions. Also, the findings point to enhancing the management of NCDs by increasing the capacity of the healthcare workforce, which is crucial. This can be achieved by providing more training opportunities for healthcare professionals and expanding the number of clinicians skilled in managing NCDs. It will be impossible to achieve global NCD targets by 2025, as part of the Sustainable Development Goals (SDGs) by 2030, without significant efforts in both policies and programs. Therefore, it is imperative to take immediate action to enhance the provision of NCD services in both public and private healthcare facilities in Nepal.

## Conclusions

The overall readiness of the facilities to provide NCDs related services in Nepal is sub-optimal. The peripheral facilities are less likely to be ready to provide NCDs related services compared to provincial and federal level hospitals. It is essential to enhance service delivery platforms and enhance the overall readiness of the health system to provide NCDs related services by increasing the number of qualified health staff, providing training, and supplying equipment and medicines.

## Data Availability

All data are publicly available in the website of DHS program (https://dhsprogram.com/data/)

## Acknowledgeme

We would like to acknowledge DHS program for providing us data for further analysis and we are grateful to those who directly or indirectly contributed us and motivated us to conduct this study.

