## Supplementary file 1 for "Non-Communicable Disease Service Readiness in Nepal: A Further Analysis of Nepal Health Facility Survey- 2021"

**Table 1: Summary of items of each domain and measurement procedure of readiness of health facilities with cardiovascular diseases related services**

| **Domain** | **Indicators** | **Measure** | **Recode** | **Calculation of score** | |
| --- | --- | --- | --- | --- | --- |
|  |  |  |  | **Domain wise score** | **Total score** |
| Guidelines and training | Guidelines (a1) | Yes | 1 | A=(a1+b1)/2*100 | (A+B+C)/3 |
|  |  | No | 0 |  |  |
|  | Staff training (b1) | Yes | 1 |  |  |
|  |  | No | 0 |  |  |
| Equipment | Stethoscope (c1) | Yes | 1 | B=(c1+d1+e1+f1+g1)/5*100 |  |
|  |  | No | 0 |  |  |
|  | Blood pressure (d1) | Yes | 1 |  |  |
|  |  | No | 0 |  |  |
|  | Adult weighing scale (e1) | Yes | 1 |  |  |
|  |  | No | 0 |  |  |
|  | Oxygen (f1) | Yes | 1 |  |  |
|  |  | No | 0 |  |  |
|  | Pulse oximeter (g1) | Yes | 1 |  |  |
|  |  | No | 0 |  |  |
| Medicines | Amlodipine/nifedipine (h1) | Yes | 1 | C=(h1+i1+j1+k1)/4*100 |  |
|  |  | No | 0 |  |  |
|  | Beta-blockers (atenolol) (i1) | Yes | 1 |  |  |
|  |  | No | 0 |  |  |
|  | Aspirin (j1) | Yes | 1 |  |  |
|  |  | No | 0 |  |  |
|  | Thiazide (k1) | Yes | 1 |  |  |
|  |  | No | 0 |  |  |

**Table 2: Summary of items of each domain and measurement procedure of readiness of health facilities with diabetes related services**

| **Domain** | **Indicators** | **Measurement** | **Recode** | **Calculation of score** | |
| --- | --- | --- | --- | --- | --- |
|  |  |  |  | **Domain wise score** | **Total score** |
| Guidelines and training | Guidelines (a2) | Yes | 1 | A=(a2+b2)/3*100 | (A+B+C)/3 |
|  |  | No | 0 |  |  |
|  | Staff training (b2) | Yes | 1 |  |  |
|  |  | No | 0 |  |  |
| Equipment | Blood pressure (c2) | Yes | 1 | B=(c2+d2+e2)/3*100 |  |
|  |  | No | 0 |  |  |
|  | Adult weighing scale (d2) | Yes | 1 |  |  |
|  |  | No | 0 |  |  |
|  | Height board/stadiometer (e2) | Yes | 1 |  |  |
|  |  | No | 0 |  |  |
| Medicines | Metformin (f2) | Yes | 1 | C=(f2+g2+h2+j2)/4*100 |  |
|  |  | No | 0 |  |  |
|  | Glibenclamide (g2) | Yes | 1 |  |  |
|  |  | No | 0 |  |  |
|  | Injectable insulin (h2) | Yes | 1 |  |  |
|  |  | No | 0 |  |  |
|  | D. Injectable glucose solution (j2) | Yes | 1 |  |  |
|  |  | No | 0 |  |  |
| Diagnostics | Blood glucose (k2) | Yes | 1 | D=(k2+I2+m2)/3*100 |  |
|  |  | No | 0 |  |  |
|  | Urine Glucose (l2) | Yes | 1 |  |  |
|  |  | No | 0 |  |  |
|  | Urine Protein (m2) | Yes | 1 |  |  |
|  |  | No | 0 |  |  |

**Table 3: Summary of items of each domain and measurement procedure of readiness of health facilities with COPD-related services**

| **Domain** | **Indicators** | **Measurement** | **Recode** | **Calculation of score** | |
| --- | --- | --- | --- | --- | --- |
|  |  |  |  | **Domain wise score** | **Domain wise score** |
| Guidelines and training | Guidelines (a3) | Yes | 1 | A=(a3+b3)/2*100 | Score=(A+B+C)/3 |
|  |  | No | 0 |  |  |
|  | Staff training (b3) | Yes | 1 |  |  |
|  |  | No | 0 |  |  |
| Equipment | Stethoscope (c3) | Yes | 1 | B=(c3+d3+e3+f3)/4*100 |  |
|  |  | No | 0 |  |  |
|  | Oxygen Flowmeter (d3) | Yes | 1 |  |  |
|  |  | No | 0 |  |  |
|  | Spacer for inhalers (e3) | Yes | 1 |  |  |
|  |  | No | 0 |  |  |
|  | Oxygen (f3) | Yes | 1 |  |  |
|  |  | No | 0 |  |  |
| Medicines | Salbutamol inhaler (g3) | Yes | 1 | C=(g3+h3+i3+j3+k3+l3+m3)/7*100 |  |
|  |  | No | 0 |  |  |
|  | Beclomethasone inhaler (h3) | Yes | 1 |  |  |
|  |  | No | 0 |  |  |
|  | Prednisolone cap/tabs (i3) | Yes | 1 |  |  |
|  |  | No | 0 |  |  |
|  | Hydrocortisone injection (j3) | Yes | 1 |  |  |
|  |  | No | 0 |  |  |
|  | Epinephrine injectable (k3) | Yes | 1 |  |  |
|  |  | No | 0 |  |  |
|  | Salbutamol inhaler (l3) | Yes | 1 |  |  |
|  |  | No | 0 |  |  |
|  | Beclomethasone inhaler (m3) | Yes | 1 |  |  |
|  |  | No | 0 |  |  |

**Table 4: Summary of items of each domain and measurement procedure of readiness of health facilities with Mental health-related services**

| **Domain** | **Indicators** | **Measurement** | **Recode** | **Calculation of score** | |
| --- | --- | --- | --- | --- | --- |
|  |  |  |  | **Domain wise score** | **Total Score** |
|  | Guidelines (a4) | Yes | 1 | A=  (a4+b4)/2*100 | (A+B)/2 |
|  |  | No | 0 |  |  |
|  | Staff training (b4) | Yes | 1 |  |  |
|  |  | No | 0 |  |  |
| Medicines | Amitriptyline (c4) | Yes | 1 | B=  (c4+d4+e4+f4+g4+h4+i4+j4)/8*100 |  |
|  |  | No | 0 |  |  |
|  | Fluoxetine (d4) | Yes | 1 |  |  |
|  |  | No | 0 |  |  |
|  | Carbamazepine (e4) | Yes | 1 |  |  |
|  |  | No | 0 |  |  |
|  | Phenobarbitone (f4) | Yes | 1 |  |  |
|  |  | No | 0 |  |  |
|  | Sodium valproate (g4) | Yes | 1 |  |  |
|  |  | No | 0 |  |  |
|  | Respiridone (h4) | Yes | 1 |  |  |
|  |  | No | 0 |  |  |
|  | Alprazolam (i4) | Yes | 1 |  |  |
|  |  | No | 0 |  |  |
|  | Diazepam (j4) | Yes | 1 |  |  |
|  |  | No | 0 |  |  |
