## Supplementary file 2 for "Non-Communicable Disease Service Readiness in Nepal: A Further Analysis of Nepal Health Facility Survey- 2021"

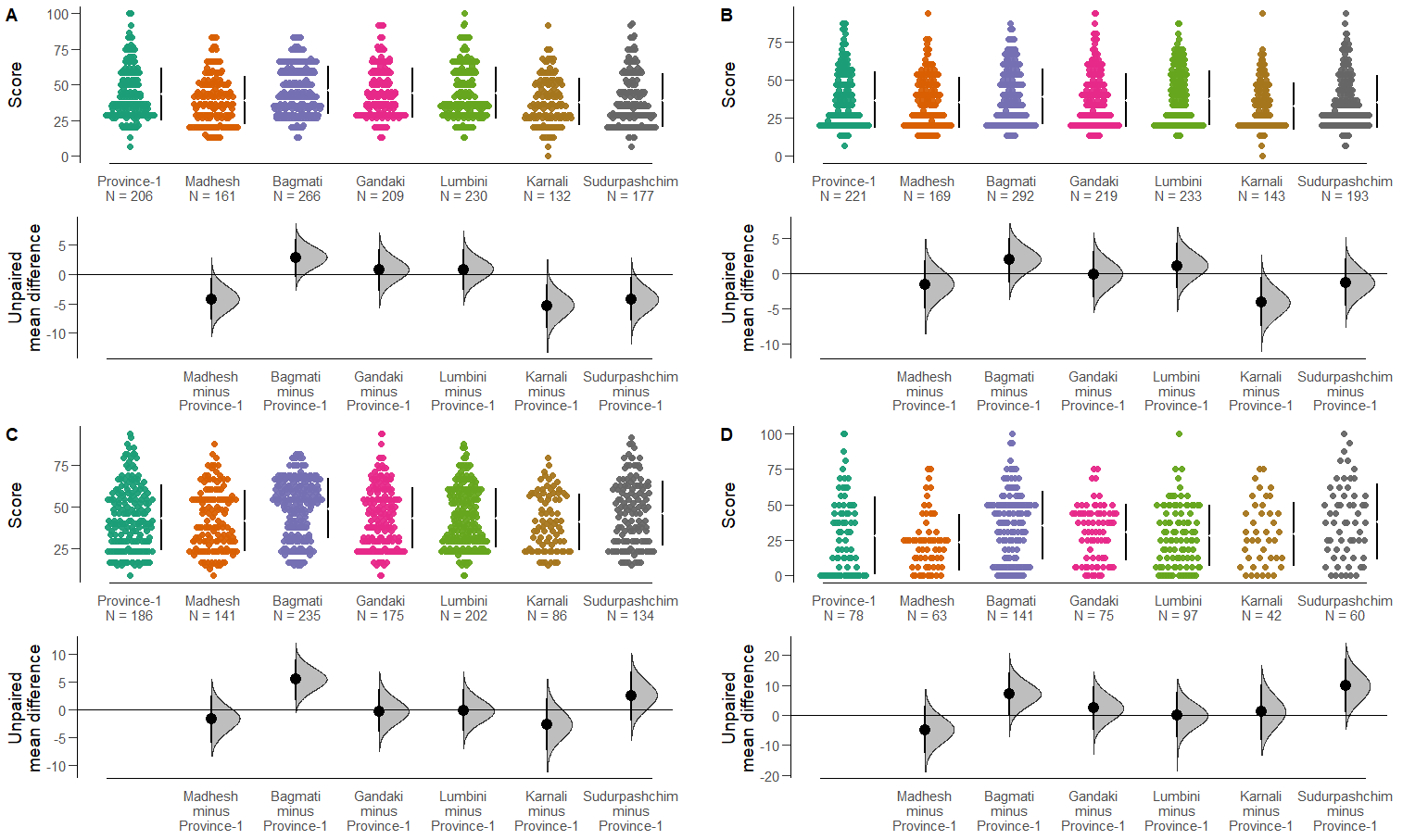

***Figure: Gardner-Altman estimation plots for comparing mean service readiness scores of facilities offering A) CVDs services, B) CRDs services, C)DM services and D) MH services, by province***
